# Real-Time Electronic Health Record Mortality Prediction During the COVID-19 Pandemic: A Prospective Cohort Study

**DOI:** 10.1101/2021.01.14.21249793

**Authors:** Peter D. Sottile, David Albers, Peter E. DeWitt, Seth Russell, J.N. Stroh, David P. Kao, Bonnie Adrian, Matthew E. Levine, Ryan Mooney, Lenny Larchick, Jean S. Kutner, Matthew K. Wynia, Jeffrey J. Glasheen, Tellen D. Bennett

## Abstract

**Background:** The SARS-CoV-2 virus has infected millions of people, overwhelming critical care resources in some regions. Many plans for rationing critical care resources during crises are based on the Sequential Organ Failure Assessment (SOFA) score. The COVID-19 pandemic created an emergent need to develop and validate a novel electronic health record (EHR)-computable tool to predict mortality.

**Research Questions:** To rapidly develop, validate, and implement a novel real-time mortality score for the COVID-19 pandemic that improves upon SOFA.

**Study Design and Methods:** We conducted a prospective cohort study of a regional health system with 12 hospitals in Colorado between March 2020 and July 2020. All patients >14 years old hospitalized during the study period without a do not resuscitate order were included. Patients were stratified by the diagnosis of COVID-19. From this cohort, we developed and validated a model using stacked generalization to predict mortality using data widely available in the EHR by combining five previously validated scores and additional novel variables reported to be associated with COVID-19-specific mortality. We compared the area under the receiver operator curve (AUROC) for the new model to the SOFA score and the Charlson Comorbidity Index.

**Results:** We prospectively analyzed 27,296 encounters, of which 1,358 (5.0%) were positive for SARS-CoV-2, 4,494 (16.5%) included intensive care unit (ICU)-level care, 1,480 (5.4%) included invasive mechanical ventilation, and 717 (2.6%) ended in death. The Charlson Comorbidity Index and SOFA scores predicted overall mortality with an AUROC of 0.72 and 0.90, respectively. Our novel score predicted overall mortality with AUROC 0.94. In the subset of patients with COVID-19, we predicted mortality with AUROC 0.90, whereas SOFA had AUROC of 0.85.

**Interpretation:** We developed and validated an accurate, in-hospital mortality prediction score in a live EHR for automatic and continuous calculation using a novel model, that improved upon SOFA.

**Take Home Points:** *Study Question:* Can we improve upon the SOFA score for real-time mortality prediction during the COVID-19 pandemic by leveraging electronic health record (EHR) data?

*Results:* We rapidly developed and implemented a novel yet SOFA-anchored mortality model across 12 hospitals and conducted a prospective cohort study of 27,296 adult hospitalizations, 1,358 (5.0%) of which were positive for SARS-CoV-2. The Charlson Comorbidity Index and SOFA scores predicted all-cause mortality with AUROCs of 0.72 and 0.90, respectively. Our novel score predicted mortality with AUROC 0.94.

*Interpretation:* A novel EHR-based mortality score can be rapidly implemented to better predict patient outcomes during an evolving pandemic.

## Introduction

The SARS-CoV-2 virus has infected >70 million and killed >1.5 million people in the year since its origination (December 2019).^1^ The resulting pandemic has overwhelmed some regions’ health care systems and critical care resources, forcing the medical community to confront the possibility of rationing resources.^2,3^ In the United States, critical care triage guidance in the setting of resource scarcity is produced at the state-level through Crisis Standards of Care (CSC) protocols.^4,5^ These protocols attempt the difficult task of ethically allocating scarce resources to individuals most likely to benefit, with the aim of saving the most lives.^6–8^ To accomplish this, CSC protocols use organ dysfunction scores and chronic comorbidity scores to assess patient survivability. Ideally, scoring would avoid systematic bias and be generalizable, accurate, flexible to circumstance, and computable within electronic health record (EHR) systems with data collected in real-time.^9^

At the foundation of most CSC protocols is the Sequential Organ Failure Assessment (SOFA) score.^10,11^ SOFA and other acuity scores, e.g., SAPSII and APACHE, are well-validated but have significant limitations. They were developed over 20 years ago before widespread electronic health records (EHRs), are rigid regarding context, and were designed to measure severity of illness and predict mortality based a few data points.^12–17^ Although SOFA predicts mortality from influenza pneumonia poorly, it was operationalized for use in patients with COVID-19.^11,18,19^ Optimizing the accuracy of mortality predictions is critical for medical triage because the decision to withhold or withdraw of life-sustaining therapies is heavily influenced by a single score in many states’ CSC protocols.^11^

The COVID-19 pandemic created an emergent need for a novel, accurate, and context-sensitive EHR-computable tool to predict mortality in hospitalized patients with and without COVID-19. Because developing a new score can take years, a predictive model must rely on well-validated scores, only adding new inputs to improve performance. Stacked generalization provides a solution.^20^ A stacked model is built upon one of or more baseline model (e.g. SOFA) and incorporates additional models only when they improve prediction.^21^

We rapidly developed and validated a novel mortality score for triage of all hospitalized patient during the COVID-19 pandemic by stacking SOFA, qSOFA, a widely used pneumonia mortality score, an acute respiratory distress syndrome (ARDS) mortality model, and a comorbidity score.^12,22–25^ We then integrated recently reported predictors that may reflect COVID-19 pathophysiology. To test the novel model, we conducted a prospective cohort study of acutely ill adults with and without COVID-19 disease.

## Study Design and Methods

We began by developing the novel mortality score using a multi-hospital retrospective cohort of 82,087 patient encounters (Figure 1 and Appendix A). We then conducted a prospective cohort study to validate the novel mortality score in patients with and without COVID-19. Our work was anchored by four goals. First, to use SOFA as a baseline and address its limitations through stacked generalization, adding other models with the potential to improve robustness and predictive performance. Second, to integrate and test potential COVID-19-specific predictors. *Third*, to rapidly deploy the new model in a live EHR across a 12-hospital system that serves more than 1.9 million patients. Fourth, to validate model performance prospectively. The Colorado Multiple Institutional Review Board approved this study (#20-0995).

**Figure 1:**
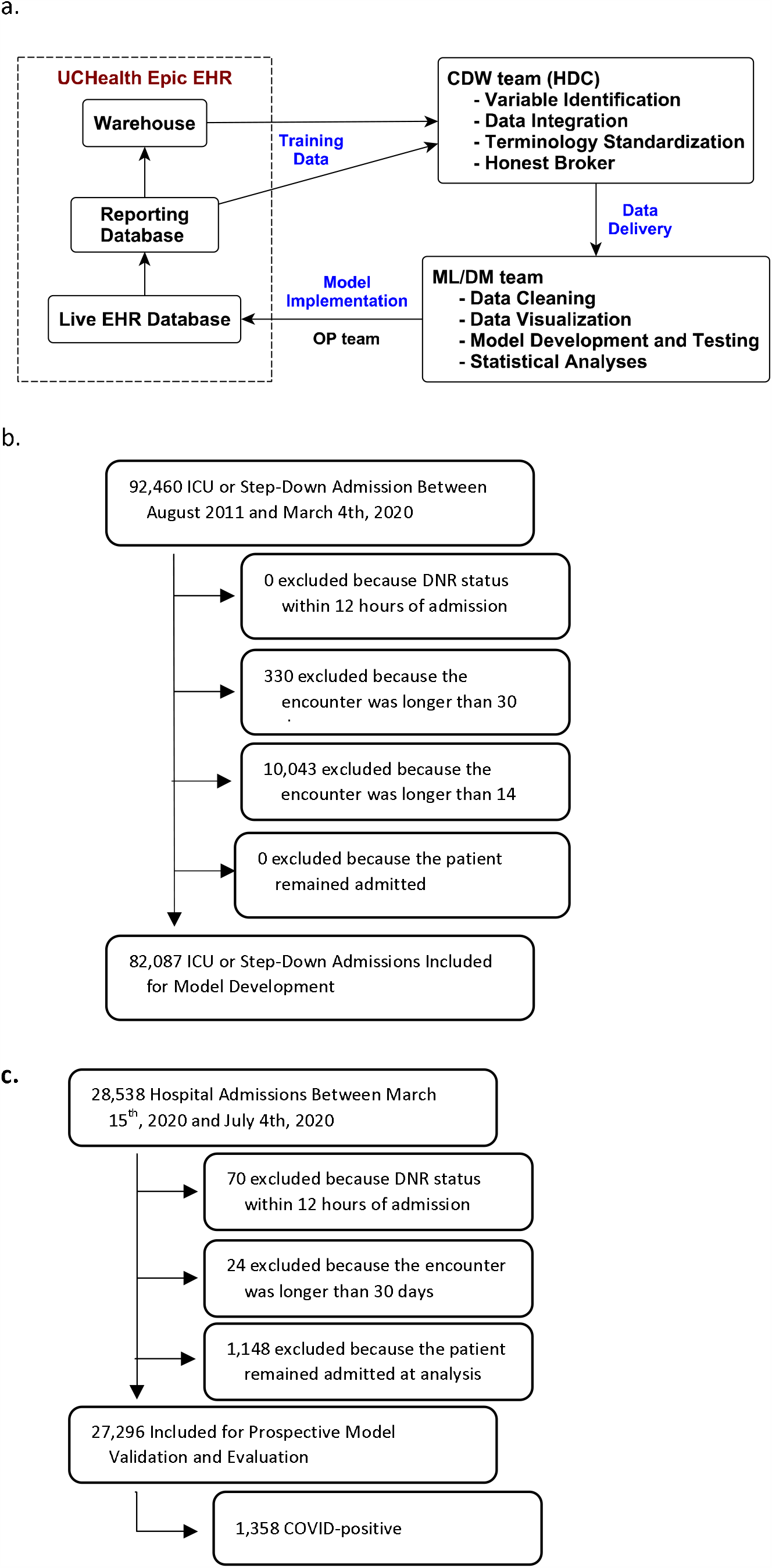
Study Data Flow: a) Data flow through the EHR and research team, b) Retrospective Cohort selection for model development, c) Prospective Cohort selection for model evaluation and validation

### Workflow and Model Deployment

Rapid development and implementation of a new score requires a full clinical and informatics pipeline including skilled data warehousing, data wrangling, machine learning, health system information technology (IT), and clinical and ethics personnel working in sync.^26–28^ All data flowed to the study team from UCHealth’s Epic instance through Health Data Compass (HDC), the enterprise data warehouse for the University of Colorado Anschutz Medical Campus (Figure 1).^29^ HDC is a multi-institutional data warehouse that links inpatient and outpatient electronic medical data, state-level all-payer claims data, and the Colorado Death Registry. The creation of data sets and models required identification of the correct data elements in both operational EHR and warehoused data tables to ensure accuracy and robustness. Rapid development, validation, and deployment of a novel model within the real-time EHR required close collaboration among three teams: 1) a data science team, 2) a clinical data warehouse team, and 3) a clinical informatics operations team (Appendix A).

This study design is consistent with recent learning health system studies.^30^ Because of the rapidly evolving pandemic, we built a data pipeline for the stacked mortality model to update as new data were captured from the EHR. We originally developed, validated and deployed the model using estimates from retrospective data, while simultaneously building technical capacity to transition to a model estimated on prospective data. The time from conception to deployment of the new model across the hospital system was one month.

### Prospective Cohort

The prospective cohort included all encounters of patients >14 years old hospitalized at any of UCHealth’s 12 acute care hospitals between March 15, 2020 (the date UCHealth halted elective procedures) through July 2020. Because CSC protocols apply to all hospitalized patients during a crisis, we included all inpatients regardless of level of care or COVID-19 status. We excluded encounters with a do not attempt resuscitation order placed within 12 hours of admission, patients who were still admitted, and encounters longer than 30 days.

### Model Methodology

We developed a model using stacked generalization to predict mortality.^20,31,32^ A stacked regression model takes other component models as covariates and estimates weights in accordance with their predictive power.^31^ We chose ridge regularized logistic regression as the top-level model to limit overfitting and to address correlation between the component models.^21^ Stacking allows for robust, accurate, and interpretable evaluation of the underlying models.^32^ Moreover, the stacked model never performs worse than the most accurate component model (see Appendix A).^33^

The stacked regression takes six logistic regression mortality models as covariates. Four are validated organ dysfunction or pneumonia/ARDS mortality prediction tools, a fifth is a comorbidity score, and a sixth is novel and COVID-specific. These models include: (1) SOFA, (2) qSOFA, (3) the CURB-65 adult pneumonia mortality score, (4) a modified version of an ARDS mortality model, and (5) a Charlson Comorbidity Index (Appendix A).^12,14,23–25^ The ARDS mortality model was attenuated to include the subset of predictors reliably available in structured form in live EHRs. The sixth model includes variables hypothesized and reported to be associated with COVID-19-specific disease severity or mortality. This includes, for example, D-dimer, lactate dehydrogenase (LDH), absolute lymphocyte count (ALC), and creatinine kinase (CK, Appendix A).^34–36^ Variables such as gender, race, or disability status were not included in any models.

### Real-time predicted mortality

Probability of mortality varies over the hospital course (Appendix B) and can be estimated at any time during the hospitalization. Thus, to estimate and validate model parameters, we selected a single time point to make a prediction – when the SOFA score reached its maximum for the encounter. Operationally, this framework allows for real-time mortality prediction under the assumption that the current measured state of the patient is the worst state the patient will experience. While this assumption will not be correct for all moments in time, it effectively underestimates the patient’s overall mortality, thus reducing the chance for premature limitation of critical care resources if used for triage decisions.

### Model Training, Evaluation, and Validation

We divided the retrospective data 40%-40%-20% for estimating the baseline logistic regression models, estimating the stacked model, and evaluating the stacked model, respectively. We estimated the stacked models with regularized (ridge) logistic regression and used 3-fold cross-validation to select a regularization parameter. The final stacked model was evaluated using empirical-bootstrap-estimated confidence intervals (CIs) and a primary metric of area under the receiver operator curve (AUROC). We validated the stacked model using the prospective cohort and the AUROC.

We chose AUROC as the accuracy metric because the primary goal of the mortality score was to generate a ranked list of patients to inform the allocation of scare resources. The AUROC is an estimate of the probability of correctly ranking a case compared to a non-case. We also estimated other accuracy metrics including positive predictive value, sensitivity, specificity, accuracy, and F1-measure (see Appendix B, eFigure1).

To evaluate the impact of COVID-19 on mortality prediction, we retrained the model using the same training strategy but limited training data to patients with COVID-19. Specifically, we divided the cohort of patients with COVID-19 40%-40%-20% for estimating the baseline logistic regression models, estimating the stacked model, and evaluating the stacked model, respectively.*Ethical Considerations*

This novel score was developed with the purpose of optimizing mortality prediction for crisis triage. Consequently, the score parameters needed to fall with the ethical framework developed for crisis triage. Briefly, in catastrophic circumstances the goal of a resource allocation processes should be to provide the most benefit to as many people as possible, and to do so in ways that sustain social cohesion and trust in the healthcare system. To maintain trust, recommendations for rationing of resources must be made prospectively, transparently and consistently across the institution and region, and by decision-makers independent of the care team. Moreover, any decision to ration resources must embrace a commitment to fairness and a proscription against rationing based on non-clinical factors such as race, gender, sexual orientation, disability, religious beliefs, citizenship status, or “VIP,” socioeconomic,or insurance status.^37–40^ Consequently, factors such a race were excluded from score development, even if they had the potential to improve accuracy.

## Results

### Cohort Characteristics and Hospital Course

The prospective cohort included a total of 28,538 encounters between March 15^th^, 2020 and July 2020. Of these, 1,148 (4.0%) were excluded because the patient remained in hospital at the time of data censoring: in-hospital survival could not be assessed. Additionally, we excluded 70 and 24 encounters respectivelydue to active DNR and encounter length>30 days. Of the remaining 27,296 encounters, 1,358 (5.0%) were positive for SARS-CoV-2, 4,494 (16.5%) included intensive care unit (ICU)-level care, 1,480 (5.4%) included invasive mechanical ventilation, and 717 (2.6%) died during the hospitalization. Of the 717 patients who received mechanical ventilation, 408 (27.6%) died. Additional demographics are in Table 1, eTable 1, and eTable 2.

**Table 1:** Prospective Cohort Characteristics and Hospital Course.

|  | **All Encounters**<br>**(N = 27,296)** | **COVID-19 Negative**<br>**(N = 25,938)** | **COVID-19 Positive**<br>**(N = 1,358)** | **P-value** |
| --- | --- | --- | --- | --- |
| **Age (SD)** | 54.3 (20.4) | 54.2 (20.5) | 56.8 (18.4) | *P* < 0.001 |
| **Female** | 15,660 (57.4%) | 15,057 (58.0%) | 603 (44.4%) | *P* < 0.001 |
| **Race** |  |  |  | *P* < 0.001 |
| **White or Caucasian** | 20,430 (74.8%) | 19,848 (76.5%) | 582 (42.9%) |  |
| **Black or African American** | 1,964 (7.2%) | 1,790 (6.9%) | 174 (12.8%) |  |
| **Other** | 4,481 (16.4%) | 3,901 (15.0%) | 580 (42.7%) |  |
| **Unknown** | 421 (1.5%) | 399 (1.5%) | 22 (1.6%) |  |
| **Ethnicity** |  |  |  | *P* < 0.001 |
| **Non-Hispanic** | 22,496 (82.4%) | 21,755 (83.9%) | 741 (54.6%) |  |
| **Hispanic** | 4,398 (16.1%) | 3,795 (14.6%) | 603 (44.4%) |  |
| **Unknown** | 402 (1.5%) | 388 (1.5%) | 14 (1.0%) |  |
| **Supplemental O2** | 16,052 (58.8%) | 14,859 (57.3%) | 1,193 (87.8%) | *P* < 0.001 |
| **High Flow Nasal Cannula** | 1,398 (5.1%) | 1,057 (4.1%) | 341 (25.1%) | *P* < 0.001 |
| **Non-Invasive Ventilation** | 1,482 (5.4%) | 1,382 (5.3%) | 100 (7.4%) | *P* < 0.001 |
| **Median Hospital Days (IQR)** | 3.0 (2.0, 5.2) | 3.0 (1.9, 5.0) | 5.5 (3.0, 9.6) | *P* < 0.001 |
| **Overall Mortality** | 717 (2.6%) | 551 (2.1%) | 166 (12.2%) | *P* < 0.001 |
| **All Mechanical Ventilation** | 1,480 (5.4%) | 1,241 (4.8%) | 239 (17.6%) | *P* < 0.001 |
| **Median Hospital Days (IQR)** | 8.4 (4.6, 15.1) | 7.7 (4.1, 13.3) | 15.2 (8.2, 21.0) | *P* < 0.001 |
| **Median ICU Days (IQR)** | 3.6 (1.6, 7.8) | 2.9 (1.4, 6.2) | 9.1 (5.3, 15.0) | *P* < 0.001 |
| **Median Ventilator Days (IQR)** | 1.8 (0.7, 5.7) | 1.4 (0.6, 3.9) | 7.5 (4.5, 12.6) | *P* < 0.001 |
| **Mortality** | 408 (27.6%) | 325 (26.2%) | 83 (34.7%) | *P* = 0.009 |

Reported p-values are to assess differences between COVID-19 Negative and COVID-19 Positive encounters.

Of the 1,358 encounters positive for COVID-19, 407 (30.0%) received ICU-level care, 239 (17.6%) were intubated, and 166 (12.2%) patients died. Of the 239 patients requiring mechanical ventilation, 83 (34.7%) died.

Compared to patients without COVID-19, patients with COVID-19 were more likely to be male (55.6% vs 42.0%, p<0.001), be Hispanic (44.4% vs 14.6%, p<0.001), receive ICU-level care (30.0% vs 15.8%, p<0.001), be intubated (17.6% vs 4.8%, p<0.001), have a longer duration of mechanical ventilation (8.7 days vs 3.0 days, p<0.001), a longer hospital length of stay (7.6 days vs 4.3 days, p<0.001), and not survive (12.2% vs 2.1%, p<0.001). Patients with COVID-19 had higher SOFA and CURB-65 scores and LDH, ferritin, and D-dimer levels than patients without COVID-19 (all p<0.05, Table 2). Mean troponin levels were lower in patients with COVID-19 compared to patients without COVID-19 (p=0.002, Table 2). However, absolute lymphocyte count and creatinine kinase levels were not dissimilar between groups (all p>0.05, Table 2).

**Table 2:** Mortality Model Inputs.

|  | All Encounters<br>(N = 27,296) | COVID-19 Negative<br>(N = 25,938) | COVID-19 Positive<br>(N = 1,358) | P-value |
| --- | --- | --- | --- | --- |
| **Scores** |  |  |  |  |
| Median qSOFA (IQR) | 0.0 (0.0, 1.0) | 0.0 (0.0, 1.0) | 0.1 (0.0, 1.0) | $P < 0.001$ |
| Median SOFA (IQR) | 2.0 (2.0, 4.0) | 2.0 (2.0, 3.0) | 3.0 (2.0, 5.0) | $P < 0.001$ |
| Median CURB-65 (IQR) | 1.0 (0.1, 2.0) | 1.0 (0.1, 2.0) | 1.0 (0.0, 2.0) | $P = 0.44$ |
| Charlson Comorbidity Index (IQR) | 1.0 (0.0, 3.0) | 1.0 (0.0, 3.0) | 1.0 (0.0, 2.0) | $P = 0.38$ |
| **ARDS Mortality Model** |  |  |  |  |
| Transfusion FFP | 59 (0.2%) | 59 (0.2%) | 0 (0.0%) | $P = 0.14$ |
| Transfusion PRBC | 396 (1.5%) | 392 (1.5%) | 4 (0.3%) | $P < 0.001$ |
| GCS $\leq 8$ | 264 (1.0%) | 246 (0.9%) | 18 (1.3%) | $P = 0.21$ |
| Lactate $> 2$ | 2,676 (9.8%) | 2,503 (9.6%) | 173 (12.7%) | $P < 0.001$ |
| Creatinine $\geq 2$ | 2,486 (9.1%) | 2,323 (9.0%) | 163 (12.0%) | $P < 0.001$ |
| Mean Bilirubin (SD) | 0.7 ± 2.0 | 0.7 ± 2.0 | 0.6 ± 0.8 | $P = 0.003$ |
| Mean Arterial pH (SD) | 7.4 ± 0.0 | 7.4 ± 0.0 | 7.4 ± 0.1 | $P = 0.001$ |
| Mean PF (SD) | 335.7 ± 212.7 | 340.7 ± 215.8 | 239.6 ± 102.0 | $P < 0.001$ |
| Mean SpO2 (SD) | 94.7 ± 2.4 | 94.7 ± 2.4 | 93.4 ± 3.1 | $P < 0.001$ |
| **Novel Predictors** |  |  |  |  |
| Mean D-Dimer (SD) | 405.0 ± 3,699.8 | 326.4 ± 2,440.3 | 1,906.2 ± 12,614.9 | $P < 0.001$ |
| Mean LDH (SD) | 229.1 ± 214.9 | 223.1 ± 207.4 | 343.5 ± 305.5 | $P < 0.001$ |
| Mean ALC (SD) | 1.4 ± 2.0 | 1.5 ± 2.0 | 1.3 ± 1.6 | $P = 0.001$ |
| Mean BUN (SD) | 19.4 ± 15.1 | 19.3 ± 14.9 | 21.2 ± 18.4 | $P < 0.001$ |
| Mean Troponin (SD) | 0.5 ± 9.0 | 0.6 ± 9.2 | 0.2 ± 3.9 | $P = 0.002$ |
| Mean CK (SD) | 173.7 ± 1,612.7 | 170.5 ± 1,567.2 | 235.4 ± 2,316.0 | $P = 0.31$ |
| Mean ALT (SD) | 21.1 ± 20.6 | 21.1 ± 21.0 | 20.9 ± 10.4 | $P = 0.47$ |
| Mean Lactate (SD) | 1.0 ± 1.1 | 1.0 ± 1.1 | 1.2 ± 1.6 | $P < 0.001$ |

The covariates included in the stacked model are calculated at a single point in time - the time of maximum SOFA score for each encounter. Presented are the summary statistics for all patients at that single point in time. FFP: fresh frozen plasma, PRBC: packed red blood cells, GCS: Glasgow coma score, PF: PaO<sub>2</sub> to FiO<sub>2</sub> ratio, LDH: lactate dehydrogenase, ALC: absolute lymphocyte count, BUN: blood urea nitrogen, CK: creatinine kinase, ALT: alanine aminotransferase

### Point-Wise Mortality Estimates

When validating mortality models in the prospective cohort, the individual component models predicted point-wise mortality (estimates of mortality risk ranging from 1-99%) with AUROCs ranging from 0.72 (Charlson Comorbidity Index) to 0.90 (SOFA) (Table 3). The stacked model predicted point-wise mortality better than any individual model: AUROC 0.94 (Figure 2). Most prospective encounters (95.7%) had predicted point-wise mortalities less than 10%. Within this group, observed mortality was only 1.0%, suggesting that the stacked model accurately identifies patients with low mortality (eTable 3).

**Table 3:** Model Area under the Receiver Operator Curve.

|  | **Retrospective Validation Cohort**<br>**(N = 16,418)** | **Prospective Validation Cohort**<br>**(N = 27,296)** | **COVID-19 Positive Validation Cohort**<br>**(N = 1,358)** |
| --- | --- | --- | --- |
| **SOFA** | 0.90 | 0.90 | 0.85 |
| **qSOFA** | 0.83 | 0.84 | 0.79 |
| **CURB-65** | 0.81 | 0.87 | 0.90 |
| **ARDS Mortality** | 0.85 | 0.88 | 0.86 |
| **Charlson Comorbidity Index** | 0.63 | 0.72 | 0.75 |
| **Novel Variables** | 0.83 | 0.88 | 0.91 |
| **Stacked Model** | 0.93 | 0.94 | 0.90 |

The AUROC for each of the component models and the final stacked model. Models were trained and validated on the initial retrospective cohort. The models were then validated on the prospective cohort and on the subset of patients with COVID-19. The AUROC for the retrospective cohort is based on a 20% holdout of the encounters for testing and evaluation. The prospective validation cohort reflects expected performance when running in a live EHR for both COVID-19 positive and negative patients.

**Figure 2:**
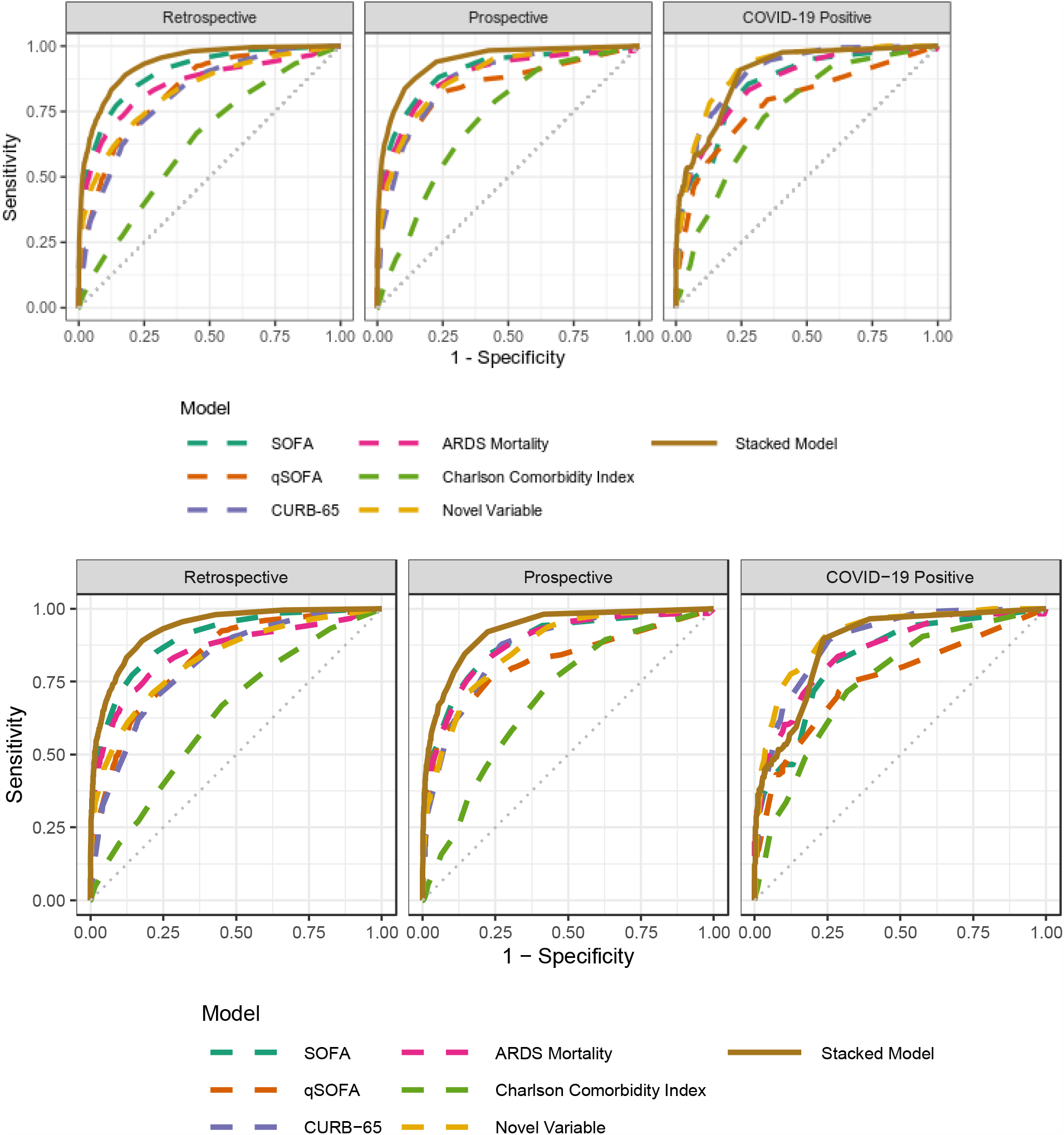
Stacked Model AUROC in the Retrospective and Prospective Cohorts: The retrospective cohort was used for training and validation (in a 40%-40%-20% split). The prospective and COVID-19 positive cohorts were used to validate the retrospectively trained model.

In patients with COVID-19, the AUROC for SOFA, CURB-65, the Charlson Comorbidity Index, and novel variables was 0.85, 0.90, 0.75, and 0.91 respectively. In this subset of patients, the stacked model predicted mortality with an AUROC of 0.90. In both analyses, the stacked model predicted mortality with narrowest 95% confidence intervals at the extremes of predicted mortality (eFigure 2). Even at moderate predicted mortalities, 95% confidence intervals were generally narrower than ten percentage points. Additional results including precision, recall, and time-integrated estimates of mortality are reported in Appendix B, eFigure 1.

When trained with retrospective data and evaluated on patients with COVID-19, the novel model outperformed the stacked model (AUROCs of 0.91 and 0.90, respectively). However, re-training the stacked model only on patients with COVID-19 improved its COVID-19-specific AUROC to 0.95 (Appendix B). The stacked model outperformed all other models for patients with COVID-19. This highlights the importance of flexible modeling constructs and suggests that patients with COVID-19 have predictors of mortality that differ from average patients.

## Discussion

We developed a new, accurate mortality prediction score that is adaptable to different diseases and settings. Improving upon SOFA and the Charlson Comorbidity Index to predict mortality, our score allows more accurate and granular ranking of patients likely to benefit from intensive care. We rapidly deployed the novel score in our EHR during the COVID-19 pandemic for potential real-time use in making triage decisions. We demonstrated that reliability was maintained in a prospective cohort of patients with and without COVID-19. Fortunately, we have not needed to use these scores for triage, but our development process forges a new path for leveraging EHRs, clinical expertise, and machine learning to provide real time, situation-critical clinical decision support.

This paper adds significantly to the literature regarding CSC and ethically allocating scarce medical resources. Like ours, most other scoring systems are based on the SOFA score, which was developed 20 years ago with simplicity and not triage in mind. SOFA predicted influenza H1N1 mortality poorly.^18,19^ Others have attempted to build novel scores that are simple and accurate.^6,7^ Our work builds on recent reports demonstrating in patients with COVID-19 that SOFA, CURB-65, PSI, APACHE II, and novel, COVID-specific COVID-GRAM scores predict mortality well: AUROC 0.87, 0.84-0.85, 0.87, 0.96, and 0.78-0.88 respectively.^41–44^ Although APACHE II out-performs other scores, it includes data that is not easily extracted from an EHR in real-time. By stacking multiple models and using data extracted in real-time from the EHR, we demonstrate similar AUROC (0.94) in a large prospective cohort of patients on whom a CSC-based triage plan would operate: those with and without COVID-19. Finally, CSC protocols have collapsed SOFA scores to rank patients in just a few categories, reflecting the difficulty of knowing when SOFA scores are sufficiently different to make a meaningful difference for triage. Our approach generates 1-99% risk of mortality and the ability to statistically differentiate between patients (or determine statistical ties) by calculating 95% CI for each score.

Our stacked model’s ability to predict mortality is tailored to our patient population in Colorado. This is important given the varied experiences with COVID-19. Our in-hospital (12% versus 21%) and ventilator mortality rates (35% versus 88%) were substantially lower than a New York cohort.^45^ Our mortality rates approach those expected for moderate-severe ARDS.^46,47^ There are potentially many explanations for these differences, including younger age, difference in comorbidities, differences in therapeutic interventions, and learning from the experience of earlier effected areas. Moreover, the utilization of ICU level of care and mechanical ventilation varies widely across the world: in New York, 14.2% of patients were treated in an ICU and 12.2% of patient received mechanical ventilation. In contrast, in a cohort of patients in China, 50.6% of patients were admitted to an ICU and 42.2% received mechanical ventilation.^35,36,41^ Such differences may affect the predictive characteristics of a mortality score. Moreover, we found that patients with COVID-19 have unique characteristics and may benefit from specific mortality prediction models. Thus, utilizing EHR data streams allows for flexibility to add additional components and retrain the stacked model as new knowledge and clinical experience accumulates. Importantly for generalization, the model can be tuned in real-time to other local patient populations and disease characteristics.

Several aspects of the informatics infrastructure and workflow are important. First, such a rapid development process would have been impossible without a robust data warehouse staffed by experts with deep knowledge of EHR data and common clinical data models. The availability of high-quality data is known to be among the largest challenges in clinical applications of machine learning.^48^ Second, our data science team was in place and had substantial shared experience with data from the health system. It would be extremely challenging to either rapidly hire or outsource the necessary expertise during a pandemic. Third, our data science team already had access to highly capable cloud-based and on-premises HIPAA-compliant computational environments. Establishing the processes and controls for such an environment takes time and expert human resources; our campus had already made those investments. Fourth, our multidisciplinary team included leadership, a variety of potential end-users, and experts from ethics, clinical informatics, machine learning, and clinical care.^26^ This diversity critically grounded the project in ethical principles and pragmatic clinical realties and allowed us to quickly iterate to a practical, implementable, and interpretable model. Because of urgent operational needs, we also had full institutional and regulatory support. Finally, we evaluated the model prospectively, an important gold-standard not often met by new machine learning-based informatic tools.^26^ Of note, there are many reports in the literature describing development of predictive models using EHR data, but very few reports of the implementation of those models in a live EHR for clinical use. In this case, the total elapsed time from including data extraction, model construction, implementation, and deployment within the EHR across the 12 UCHealth hospitals was 1 month, illustrating the potential capacity for novel predictive model development. Now that we have demonstrated a workflow to rapidly develop new informatics tools in our health system, we anticipate that many other tools will follow.

This manuscript has several limitations. First, all scores are calculated from EHR data. While this allows for real-time score calculation, it introduces the possibility of artifactual data skewing mortality prediction. This was partially addressed by placing acceptable ranges on physiologic variables (see Appendix A). Second, missing data or data collected at different time intervals is inherent in the analysis of EHR data. To overcome this, we developed a system of imputation and last known value carry forward (see Appendix A). Such assumptions may introduce systematic and unmeasured bias but are unavoidable operationally. Third, more sophisticated machine learning techniques—e.g., Gaussian process regressions—may allow for more accurate mortality predictions.^49^ However, we chose methods that were robustly estimable and would allow for transparent interpretation of underlying model contributions to the overall score. Fourth, in-hospital mortality may not be the optimal metric to make triage decisions. One-year mortality may be a better metric but, given the desire to validate a mortality predictor quickly, longer-term outcomes were not available. Fifth, our data and patient population are specific to Colorado and results may differ geographically. Finally, some clinical indicators of illness severity were not included in the models, e.g. prone positioning, continuous renal replacement therapy, and radiographic results. These data may improve mortality prediction but are difficult to routinely and reliably auto-extract from the EHR.

## Conclusion

We developed a novel and accurate in-hospital mortality score that was deployed in a live EHR and automatically and continuously calculated for real-time evaluation of patient mortality. The score can be tuned to a local population and updated to reflect emerging knowledge regarding COVID-19. Moreover, this score adheres to the ethical principles necessary for triaging.^37–40^ Further research to test multi-center score performance, refine mortality prediction over longer periods of time, and investigate the optimal methods to use such a score in a CSC protocol is needed.

## Data Availability

The data referred to in the manuscript are the property of UCHealth and we are unable to share them.

## Abbreviations

ALC: absolute lymphocyte count
APACHE II: Acute Physiology and Chronic Health Evaluation
ARDS: acute respiratory distress syndrome
AUROC: area under the receiver operating curve
CCI: Charlson Comorbidity Index
CI: confidence interval
CK: creatinine kinase
CSC: crisis standard of care
EHR: electronic health record
HDC: Health Data Compass
ICU: intensive care unit
IT: information technology
LDH: lactate dehydrogenase
SAPS II: Simplified Acute Physiology Score
SOFA: Sequential Organ Failure Assessment
PSI: Pneumonia Severity Index

## Acknowledgements

Guarantor: Tellen D. Bennett

## Author Contributions

TDB had full access to all of the data in the study and takes responsibility for the integrity of the data and the accuracy of the data analysis. PDS, DA, PED, SR, JS, TDB contributed substantially to the study design, data acquisition, data analysis and interpretation, and the writing of the manuscript. DPK, BA, RM, and LL contributed substantially to data acquisition, verifying data integrity, and the writing of the manuscript. MEK contributed substantially to study design and the writing of the manuscript. JSK, MKW, JJG contributed substantially to the study design, data acquisition, and the writing of the manuscript.

## Other Contributions

We would like to acknowledge Sarah Davis, Michelle Edelmann, and Michael Kahn at Health Data Compass.

