## Supplemental Appendix for "Real-Time Electronic Health Record Mortality Prediction During the COVID-19 Pandemic: A Prospective Cohort Study"

**Appendix Table of Contents:**

**Appendix A: Additional Methods**

Team Roles

Component Models

Variable Definitions

Missing Data, Repeated Data, and Outliers

Time-Integrated Mortality Prediction

Statistical Analysis and Software

**Appendix B: Additional Results**

Retrospective Cohort for Initial Model Training

Confusion Matrix Assessments

Time-Integrated Mortality Estimates

Models Trained on COVID-19 Specific Data

**Appendix C: References**

**eTable 1: Retrospective versus Prospective Cohort Demographic Characteristics and Hospital Course**

**eTable 2: Mortality Model Inputs**

**eTable 3: Actual Mortality by Predicted Mortality Decile in the Prospective Cohort**

**eTable 4: Stacked Model Coefficients in the Retrospective, Prospective, and COVID-19 Cohorts**

**eTable 5: Mortality Prediction 3 and 7 Days After Admission**

**eTable 6 and eFigure 1: Confusion Matrix**

**eFigure 2: Confidence Intervals of Point-Wise Predicted Mortality in the Retrospective and Prospective Cohorts**

**eFigure 3: Average Predicted Mortality Stratified by Actual Mortality over the course of the Hospitalization in the Retrospective and Prospective Cohorts**

**Appendix A: Additional Methods**

*Team Roles*

Rapid development, validation, and deployments of a novel model within the real-time EHR required cross-functional team collaboration among three teams: 1) a data science team, 2) a clinical data warehouse team (CDW), and 3) a clinical informatics operations team (OPS). Clinical expertise was present on all three teams and some individuals were present on multiple teams. The CDW and OPS teams worked to identify the clinical variables necessary. The CDW team constructed the retrospective and prospective data sets. The creation of the data sets and models required both the operational data and the CDW data to identify the correct data elements to ensure accuracy and robustness. The DS team constructed and validated the models. The DS and OPS teams worked to integrate the model within the operational EHR. All three teams were required for model validation within the operational system.

*Component Models*

The following variables were included in each component model:

1. SOFA: PaO2 to FiO2 ratio (PF), mechanical ventilation, platelets, Glasgow coma scale (GCS), bilirubin, mean arterial pressure (MAP), vasoactive medications, creatinine
2. qSOFA: GCS, systolic blood pressure (SBP), respiratory rate (RR)
3. CURB-65: age, systolic and diastolic blood pressure (SBP and DBP), blood urea nitrogen (BUN), Glasgow coma scale (GCS), and respiratory rate
4. ARDS Mortality Score: This was limited to a subset of variables available in structured form in the EHR. This includes arterial pH, bilirubin, PF ratio, SpO2, mechanical ventilation, transfusion of fresh frozen plasma (FFP), transfusion of red blood cells (PRBC), GCS, shock (defined as lactate >2.0) and acute kidney injury (defined as a creatinine >= 2.0).
5. Charlson Comorbidity Index (CCI): For retrospective data, this was calculated using ICD-9 and ICD-10 data available from previous encounters in the EHR. The open source R package ICD was used to transform these codes into a final Charlson Index.^1^ In the live EHR, ICD-9 and ICD-10 codes are not available in real-time during an active encounter. Consequently, the health system developed and implemented a system to capture CCI data on all new admissions. Mining historical records for data relevant to the CCI calculation was possible, but bias against patients with a long history at our institution compared to patients who have never been treated at UCHealth.
6. Novel Variables: age (continuous), absolute lymphocyte count (ALC), alanine aminotransferase (ALT), brain natriuretic peptide (BNP), BUN, creatinine kinase (CK), D-dimer, lactate, lactate dehydrogenase (LDH), and troponin. Ferritin and CRP were not included as novel variables, despite data suggesting that they may predict outcomes in COVID-19 because too many patients were lacking data.

*Variable Definitions*

1. All vital sign, medication, and laboratory variables were directly extracted from the EHR. If not directly recorded in the EHR, MAP was calculated from SBP and DBP.
2. Mechanical Ventilation: Defined as the presence of an endotracheal tube or tracheostomy in the Line, Drain, Airway (LDA) table from the EHR
   a. Placement date-time set to admission date-time if missing or before admission
   b. Removal date-time set to discharge date-time if missing
   c. Overlapping ventilation episodes were collapsed into one episode, e.g., the temporal order of tube 1 placed > tube 2 placed > tube 1 removed > tube 2 removed was formatted into a single episode starting with the placement of tube 1 and concluding with the removal of tube 2.
   d. Distinct events with less than two hours lapsing between removal of initial tube and placement of second tube were collapsed into a single mechanical ventilation episode.
3. PaO2 to FiO2 Ratios (PF ratio): PF ratio was calculated in a stepwise fashion. First, if the PaO2 and FiO2 were both known, it was calculated directly. This occurred if a patient had a recent ABG and was on invasive mechanical ventilation, non-invasive positive pressure ventilation, or heated high flow nasal cannula – all modes where FiO2 is set directly. Second, if the patient was on nasal cannula or face mask, the FiO2 needed to be estimated based on flow rate (see below). Third, if the PaO2 was not directly measured, the PF ratio was estimated from the SpO2 to FiO2 ratio as described elsewhere.^2–5^ For SpO2 and FiO2 outside provided ranges, the following logic was used: If FiO2 < 30 and SpO2 between 80 and 96, set PF ratio to '399' which results in a 1 for SOFA respiratory score. If FiO2 < 30 and SpO2 > 96, set PF ratio to '401' which results in a 0 for SOFA respiratory score. Finally, if the imputed SpO2 to FiO2 could not be imputed, the PaO2 was estimated from the FiO2 using the alveolar gas equation (assuming atmospheric pressure of 620, pCO2=40, R=0.8, and A-a gradient of 10.

*Missing Data, Repeated Data, and Outliers*

Vital signs, labs, and other covariates are collected throughout the encounter. The date and time of known values that a given covariate was measured was used to construct a time series of data. Because the collection of information is not synchronized in time, at any given time, many of the covariates are missing data. For example, a PaO2 taken at 13:01:00 and respiratory rate taken at 13:04:00 would result in the PaO2 value missing in the row for time 13:04:00 and respiratory rate missing in the row for time 13:01:00.

Four solutions are commonly utilized. The first is to impute the linear interpolation between known values. The second is to use the last known observation within a specific window of time (last observation carried forward, LOCF). The third is to use the next observation within a specified window of time (next observation carried backward, NOCB). The fourth is to use a 'normal’ default if no values were present during the entire hospitalization and it could not otherwise be estimated. This explicitly assumes that if a value was not checked, it was because the clinical team did not think it was going to be sufficiently different from normal to change management. We did the following:

*Indicator variables:* LOCF as appropriate for temporal associations, e.g., actively intubated, or set for the entire encounter record if no temporal considerations were needed.

*Vital Signs* (SBP, DPB, MAP, RR, Spo2): NOCB was utilized until the time of the first measured, linear interpolation was utilized between serial measurements, and LOCF from the last measurement to the end of the encounter.

*Medications*: LOCF was used for all medication doses.

*Bilirubin, BUN, Platelets, and Lactate*: Linear interpolation was utilized between subsequent values.

*D*-*dimer, ALC, troponin, BNP, CK, arterial pH:* NOCB was utilized until the time of the first measured, linear interpolation was utilized between serial measurements, and LOCF from the last measurement to the end of the encounter.

*GCS*: GCS required special attention. It is commonly obtained in the emergency department, but in our health system is less likely to be used once the patient is intubated. Consequently, we used LOCF within a 24h window if it was recorded. If the patient was intubated, GCS was assumed to be 11. Otherwise, GCS was assumed to be 15.

*Normal values:*

1. PaO2 = 540 (mmHg)
2. FiO2 = 0.21
3. SpO2 = 95
4. Platelet = 250 (10^9^ /L)
5. Bilirubin = 0.2 (mg / dL)
6. BUN = 14 mg/dL
7. Creatinine = 0.75
8. Dobutamine = 0 (mcg / kg / min)
9. Epinephrine = 0 (mcg / kg / min)
10. Norepinephrine = 0 (mcg / kg / min)
11. D-dimer = 250 (ng / mL)
12. LDH = 210 (U / L)
13. Absolute Lymphocyte Count = 1.4 (10^9^ /L)
14. Troponin = 0.02 (ng / mL)
15. BNP = 100 (pg / mL)
16. Lactate = 0.75 (mmol / L)
17. CK = 110 (U / L)
18. ALT = 20 (U / L)
19. Arterial pH = 7.40

Sometimes multiple values for the same variable were available in the EHR at the same instance in time. In this case, different logic was applied to the variable in question to reduce values to one observation.

1. *PaO2, FiO2, SpO2:* For a given instant of time, keep largest value. If an FiO2 was present at the same time as a L/min flowrate, the FiO2 value was used.
2. *Absolute Lymphocyte Count, ALT, Arterial pH, Bilirubin, BNP, BUN, CK, Creatinine, D-dimer, Lactate, LDH, Platelet count, Troponin:* For a given instant of time, keep lab with first result time (most labs used the time the lab was obtained)
3. *Blood Pressure:* For timepoints with multiple SBP, DBP, and MAP values, keep value with highest MAP.
4. *Respiratory Rate:* Keep “Actual RR” over “Observed RR”; otherwise, keep highest value.
5. *GCS*: Prefer MD stated over other values; keep largest value if still duplicates
6. *COVID Status:* If multiple values present, prefer 'Positive' first, if 'Positive' not present, then prefer 'Negative'.
7. *Medications:* Dose (mainly drip rates) were tracked in 15-minute increments to avoid rapid fluctuations when changing bags.

Outliers were observed primarily in the vital sign data. For instance: a respiratory rate of 120 is likely 12 or 20; a MAP of 350 is likely a measurement being recorded while an arterial catheter is being flushed. To account for this, we set reasonable ranges on the values for vital signs. Measurements outside this range were clipped to the minimum or maximum value.

1. Respiratory Rate: Values below 0 or above 85 were discarded. Remaining values were clipped to the range of 4 to 60
2. Systolic Blood Pressure: Values below 11 or above 350 were discarded. Remaining values were clipped to the range of 30 to 300
3. Diastolic Blood Pressure: Values below 10 were discarded. Palpation values were set to 20. Remaining values were clipped to the range of 20 to 250
4. Mean Arterial Blood Pressure: MAP is normally machine reported. If missing or if MAP above SBP and below DBP, MAP was calculated according to the formula (SBP + 2 * DBP) / 3. Remaining values were clipped to the range 20 to 250
5. SpO2: 0 to 100
6. FIO2: Acceptable rage 0.21 to 1.0. Additionally, for patients on nasal cannula or simple face mask: if O2 flow rate above 15 L/min, cap at 15 L/min. Where FiO2 was missing but L/min rate present, used the formula$21+(4*Flow Rate)$ to convert. Invalid flow rates (negative values, above 100) were first discarded. If Nasal cannula FiO2 (either reported or calculated) above 60%, cap to 60%.
7. GCS: integer values between 3 and 15
8. Vasopressor medications: mcg/kg/min above 100 were dropped.

*Time-integrated mortality prediction*:

From a crisis triage standpoint, all critical care interventions are considered a time limited trial. Therefore, in addition to an initially estimate of mortality, a score to facilitate reassessment and ongoing utility of any intervention is necessary. Consequently, we also predicted mortality after three and seven days of hospitalization. This allowed us to incorporate change in the patient condition over time—the acuity trajectory—as a predictor of mortality, rather than simply utilizing physiologic data from a single point in time. To incorporate the acuity trajectory, we estimated the point-wise mortality score every time a covariate was updated during the patient encounter, creating a time series of predicted mortalities. We then calculated seven parameters and used them as covariates in a stacked ridge logistic regression predicting mortality. These included the initial and maximum SOFA score over the timespan of interest, as these measures have been shown to predict mortality over a patients’ hospitalization.^6^ Second, we estimated the integral of the stack-model-estimated point-wise mortality over time. Third, we estimated the linear trend of point-wise predicted mortality over time with linear regression and included both the slope and the standard error of the slope. Finally, we included both the first point-wise predicted mortality and the maximum point-wise predicted mortality over the time span of interest.

*Statistical Analysis and Software*

We performed univariable comparisons of the demographic and baseline characteristics of the retrospective and prospective cohorts and between patients with and without COVID-19 in the prospective cohort using chi-square tests for categorical data and unpaired t-tests for continuous variables. All analyses were performed using R version 3.6.^7^

**Appendix B: Additional Results**

*Retrospective Cohort for Initial Model Training*

The retrospective cohort included all encounters of patients >14 years old hospitalized at any of UCHealth’s 12 hospitals between August 2011 and March 4, 2020 whose hospital stay included admission to either an intensive care unit (ICU) or intermediate care unit. We restricted the retrospective data to encounters completed before March 5, 2020, the date of the first reported COVID-19 case in Colorado.^8^ We excluded encounters with a do not attempt resuscitation order placed within 12 hours of admission or a duration exceeding 14 days, as mortality after prolonged hospitalization likely represents different physiology than mortality from an acute event.

The retrospective cohort included 82,087 encounters by 63,290 unique patients. Of these encounters, 59,733 (72.8%) required ICU-level care, 14,847 (18.1%) required invasive mechanical ventilation, and 5,726 (7.0%) ended in death. Patients had an average age of 58.1 ± 17.8 years and 35,826 (43.6%) were female. Demographics, clinical characteristics, and hospital course are shown in eTable 1. Model inputs are shown in eTable 2.

Because the prospective cohort included all admissions and not just ICU or intermediate care admissions, patients in the prospective cohort were less likely to receive ICU level-of-care (16.5% vs 72.8%, p<0.0001), less likely to be intubated (5.4% vs 18.1%, p < 0.0001), and less likely to die (2.6% vs 7.0% vs, p<0.0001) when compared to patients in the retrospective cohort (eTable 1).

*Confusion Matrix Assessments*

The intended use of the model is to rank patients for triage. As such, the most important metric is the relative ranking of cases (encounters ending in expiration of the patient) and non-cases (encounters concluding with the patient discharged alive). If we were to randomly select an encounter ending in death and an encounter ending in survival, then our model should provide a higher probability of mortality estimate for the former than the latter. The value of the estimated probability is less relevant for this intended use. If the encounter ending in mortality had an estimated probability of mortality of 0.08 and the encounter ending in survival had an estimated probability of mortality of 0.05, then our model is correctly ranking patients. This would also be true if the respective probability estimates were 0.99 and 0.90. The AUROC is a measure of the probability of correctly ranking a case and non-case.

A health system might choose to activate a clinical decision support alert at an estimated mortality probability. Assessing the model in correctly discriminating between encounters ending in mortality/survival is dependent on arbitrary thresholds to turn the continuous probability of mortality estimate to a binary survived/expired. eFigure 1 and eTable 6 shows common confusion matrix statistics as a function of the threshold.

*Time-Integrated Mortality Estimates:*

A smoothed average probability of mortality over the course of the hospitalization and stratified by disposition shows that on average, encounters ending in the patient’s expiration will have probability of mortality estimates increasing and decreasing over time (eFigure 3). Moreover, for patients who expire the estimated probability of mortality increases within the final hours of the encounter. Encounters where the patient is discharged alive tend to have, on average, a lower probability of mortality and a relatively smooth trajectory.

Despite this limitation, models predicting mortality at 3 and 7 days after admission performed relatively well, although worse than the point-wise stacked models: AUROC 0.84 (3 days) 0.82 (7 days) in the retrospective cohort. Validation of these models in the prospective cohort and COVID-19 positive patients, mortality prediction accuracy was only moderate: AUROC 0.83 and 0.77 (3 days) and 0.84 and 0.80 at 7-days, respectively (eTable 4).

*Models Trained on COVID-19 Specific Data*

To understand how patients with COVD-19 may differ from the population as whole, we retrained the stacked model to predict point-wise mortality using patients in the prospective cohort and on patients with COVID-19 only. We divided the COVID-19 positive patient data 40%-40%-20% for training the logistic regression models, training the stacked model, and evaluating the stacked model, respectively. We estimated the stacked models with regularized (ridge) logistic regression and used 3-fold cross-validation to select a regularization parameter. The final stacked model was evaluated using bootstrap-estimated confidence intervals (CIs) and a primary metric of area under the receiver operator curve (AUROC). These cohorts are substantially smaller than the retrospective cohort, and care must be used when interpreting AUROC results.

However, we noted some qualitative differences in the weights given to different models between training groups. In general, SOFA and the ARDS mortality model received similar weights across all three cohorts, suggesting it remains a robust predictor of mortality. QSOFA was weighted substantially more, while CURB-65 and the Charlson Comorbidity Index were all weighted substantially less when trained with COVID-19 only cohorts. Interestingly, the novel variables were also weighed less in the COVID-19 only cohort (eTable 5). We hypothesize that these weights change between training cohorts because the novel variables and SOFA becomes more closely correlated in patients with COVID-19 than in patients without COVID-19.

In patients with COVID-19, models trained on COVID-19 patients had an AUROC for SOFA, CURB-65, the Charlson Comorbidity Index, and novel variables of 0.86, 0.90, 0.75, and 0.89 respectively. In this subset of patients, the stacked model predicted mortality with a AUROC of 0.95. The stacked model continued to predict mortality with narrowest 95% confidence intervals at the extremes of predicted mortality but given the smaller sample size at moderate predicted mortalities, 95% confidence intervals had a width of less of thirty percentage points (data not shown).

**Appendix C: References**

7.	R Core Team. R: A language and environment for statistical computing. 2017.

8.	Health CD of P. Colorado Deparment of Public Health: COVID-19 Updates. https://covid19.colorado.gov/data/case-data. Accessed May 15, 2020.

**eTable 1: Retrospective versus Prospective Cohort Demographic Characteristics and Hospital Course**

|  | Retrospective Encounters  (N = 82,087) | Prospective Encounters  (N = 27,296) | P-value |
| --- | --- | --- | --- |
| Age (SD) | 58.1 (17.8) | 54.3 (20.4) | *P* < 0.001 |
| Female | 35,826 (43.6%) | 15,660 (57.4%) | *P* < 0.001 |
| Race |  |  | *P* < 0.001 |
| White or Caucasian | 58,915 (71.8%) | 20,430 (74.8%) |  |
| Black or African American | 8,651 (10.5%) | 1,964 (7.2%) |  |
| Other | 12,763 (15.5%) | 4,481 (16.4%) |  |
| Unknown | 1,758 (2.1%) | 421 (1.5%) |  |
| Ethnicity |  |  | *P* < 0.001 |
| Non-Hispanic | 69,772 (85.0%) | 22,496 (82.4%) |  |
| Hispanic | 10,489 (12.8%) | 4,398 (16.1%) |  |
| Unknown | 1,826 (2.2%) | 402 (1.5%) |  |
| Supplemental O2 | 66,013 (80.4%) | 16,052 (58.8%) | *P* < 0.001 |
| High Flow Nasal Cannula | 5,729 (7.0%) | 1,398 (5.1%) | *P* < 0.001 |
| Non-Invasive Ventilation | 5,925 (7.2%) | 1,482 (5.4%) | *P* < 0.001 |
| Median Hospital Days  (IQR) | 4.04 [2.2, 7.0] | 3.0 (2.0, 5.2) | *P* < 0.001 |
| Overall Mortality | 5,726 (7%) | 717 (2.6%) | *P* < 0.001 |
| All Mechanical Ventilation | 14,847 (18.1%) | 1,480 (5.4%) | *P* < 0.001 |
| Median Hospital Days  (IQR) | 6.3 (3.3, 9.3) | 8.4 (4.6, 15.1) | *P* < 0.001 |
| Median ICU Days (IQR) | 2.3 (1.1, 4.3) | 3.6 (1.6, 7.8) | *P* < 0.001 |
| Median Ventilator Days  (IQR) | 1.0 (0.5, 2.3) | 1.8 (0.7, 5.7) | *P* < 0.001 |
| Mortality | 4,055 (27.3%) | 408 (27.6%) | *P* = 0.86 |

**eTable 2: Mortality Model Inputs**

|  | Retrospective Encounters  (N = 82,087) | Prospective Encounter  (N = 27,296) | P-value |
| --- | --- | --- | --- |
| Scores |  |  |  |
| Median qSOFA (IQR) | 0.9 [0.0. 1.0] | 0.0 [0.0, 1.0] | *P* < 0.001 |
| Median SOFA (IQR) | 3.00 [2.0, 6.0] | 2.0 [2.0, 4.0] | *P* < 0.001 |
| Median CURB-65  (IQR) | 1.6 [1.0, 2.2] | 1.0 [0.1, 2.0] | *P* < 0.001 |
| Charlson  Comorbidity  Index (IQR) | 2.0 [1.0, 4.0] | 1.0 [0.0, 3.0] | *P* < 0.001 |
| ARDS Mortality Model |  |  |  |
| Transfusion FFP | 264 (0.3%) | 59 (0.2%) | *P* = 0.007 |
| Transfusion PRBC | 853 (1.0%) | 396 (1.5%) | *P* < 0.001 |
| GCS ≤ 8 | 2,676 (3.3%) | 264 (1.0%) | *P* < 0.001 |
| Lactate > 2 | 12,428 (15.1%) | 2,676 (9.8%) | *P* < 0.001 |
| Creatinine ≥ 2 | 11,860 (14.4%) | 2,486 (9.1%) | *P* < 0.001 |
| Mean Bilirubin (SD) | 0.8 ± 2.4 | 0.7 ± 2.0 | *P* < 0.001 |
| Mean Arterial pH  (SD) | 7.4 ± 0.1 | 7.4 ± 0.0 | *P* < 0.001 |
| Mean PF (SD) | 426.6 ± 575.9 | 335.7 ± 212.7 | *P* < 0.001 |
| Mean SpO2 (SD) | 94.7 ± 3.2 | 94.7 ± 2.4 | *P* = 0.74 |
| Novel Predictors |  |  |  |
| Mean D-Dimer (SD) | 391.9 ± 3,075.1 | 405.0 ± 3,699.8 | *P* = 0.60 |
| Mean LDH (SD) | 222.7 ± 221.6 | 229.1 ± 214.9 | *P* < 0.001 |
| Mean ALC (SD) | 1.4 ± 3.3 | 1.4 ± 2.0 | *P* = 0.81 |
| Mean BUN (SD) | 22.3 ± 18.1 | 19.4 ± 15.1 | *P* < 0.001 |
| Mean Troponin (SD) | 1.8 ± 15.6 | 0.5 ± 9.0 | *P* < 0.001 |
| Mean CK (SD) | 195.4 ± 2,240.9 | 173.7 ± 1,612.7 | *P* = 0.08 |
| Mean ALT (SD) | 23.2 ± 73.2 | 21.1 ± 20.6 | *P* < 0.001 |
| Mean Lactate (SD) | 1.3 ± 1.9 | 1.0 ± 1.1 | *P* < 0.001 |

The covariates included in the stacked model at the time the maximum SOFA score for each encounter. FFP: fresh frozen plasm, PRBC: packed red blood cells, GCS: Glasgow comas score, PF: PaO2 to FiO2 ratio, LDH: lactate dehydrogenase, ALC: absolute lymphocyte count, BUN: blood urea nitrogen, CK: creatinine kinase, ALT: alanine aminotransferase

**eTable 3: Actual Mortality by Predicted Mortality Decile in the Prospective Cohort**

|  | |  | | Retrospective | | Prospective | | COVID-19 Positive | | |
| --- | --- | --- | --- | --- | --- | --- | --- | --- | --- | --- |
|  | | Predicted Mortality | | **Encounters** | **Expired** | **Encounters** | **Expired** | **Encounters** | | Expired |
|  | | <10% | | 14,598 (88.9%) | 329 (2.3%) | 26,118 (95.7%) | 269 (1.0%) | 1,160 (85.4%) | | 69 (5.9%) |
|  | | 11 - 20% | | 638 (3.9%) | 139 (21.8%) | 442 (1.6%) | 72 (16.3%) | 64 (4.7%) | | 10 (15.6%) |
|  | | 21 - 30% | | 260 (1.6%) | 62 (23.8%) | 172 (0.6%) | 46 (26.7%) | 22 (1.6%) | | 10 (45.5%) |
|  | | 31 - 40% | | 137 (0.8%) | 55 (40.1%) | 116 (0.4%) | 38 (32.8%) | 15 (1.1%) | | 5 (33.3%) |
|  | | 41 - 50% | | 104 (0.6%) | 56 (53.8%) | 72 (0.3%) | 30 (41.7%) | 18 (1.3%) | | 7 (38.9%) |
|  | | 51 - 60% | | 88 (0.5%) | 50 (56.8%) | 63 (0.2%) | 24 (38.1%) | 14 (1.0%) | | 9 (64.3%) |
|  | | 61 - 70% | | 94 (0.6%) | 56 (59.6%) | 46 (0.2%) | 26 (56.5%) | 7 (0.5%) | | 4 (57.1%) |
|  | | 71 - 80% | | 99 (0.6%) | 62 (62.6%) | 50 (0.2%) | 36 (72.0%) | 13 (1.0%) | | 10 (76.9%) |
|  | | 81 - 90% | | 114 (0.7%) | 81 (71.1%) | 67 (0.2%) | 42 (62.7%) | 13 (1.0%) | | 10 (76.9%) |
| 91 - 100% | 286 (1.7%) | | 264 (92.3%) | 150 (0.5%) | 134 (89.3%) | 32 (2.4%) | 32 (100.0%) | |  |  |

The percentages reported for encounters are column percentages, i.e., percentage of all encounters. The percentages for expired are row percentages, i.e., the percentage of the encounters noted in the row which concluded in the patient expiring.

**eTable 4: Mortality Prediction 3 and 7 Days After Admission**

|  | Retrospective | Prospective | COVID-19 Positive |
| --- | --- | --- | --- |
| First 3 days | 0.84 | 0.83 | 0.77 |
| First 7 days | 0.82 | 0.84 | 0.80 |

Table cells shows AUROC in each cohort at time point. The point-wise mortality scores were calculated at all time points with complete data. Using this time series, a logistic regression model using several features from the mortality time-series was trained and validated with the retrospective cohort (in a 40%-40%-20% split). The prospective and COVID-19 positive cohorts were used to validate the retrospectively trained logistic regression model.

**eTable 5: Stacked Model Coefficients in the Retrospective, Prospective, and COVID-19 Cohorts**

|  | Retrospective Trained | Prospective Trained | COVID-19 Trained |
| --- | --- | --- | --- |
| SOFA | 1.22 | 1.76 | 0.86 |
| qSOFA | -0.10 | -2.45 | -1.41 |
| CURB-65 | 1.22 | 3.02 | 2.73 |
| ARDS Mortality | 2.63 | 2.96 | 2.19 |
| Charlson Comorbidity Index | 4.64 | 9.21 | 4.18 |
| Novel | 2.20 | 3.74 | 2.47 |

Coefficients of the models in log-odds

**eTable 6 and eFigure 1: Confusion Matrix**

The purpose of the main stacked model was to create a ranked patient list by probability of mortality. If the model was to be used to as part of a clinical decision support alert, then a threshold for the estimated probability would need to be used to define when an alert fires.

For example, if the threshold was set at 0.40, then the reported models have the following performance statistics:

|  | Positive Predictive Value | Sensitivity | Specificity | Accuracy | F1 |
| --- | --- | --- | --- | --- | --- |
| Retrospective | 0.72 | 0.49 | 0.99 | 0.95 | 0.59 |
| Prospective | 0.65 | 0.41 | 0.99 | 0.98 | 0.50 |
| COVID-19 Positive | 0.79 | 0.43 | 0.98 | 0.91 | 0.55 |

The figure below shows performance metrics across all potential probability thresholds.

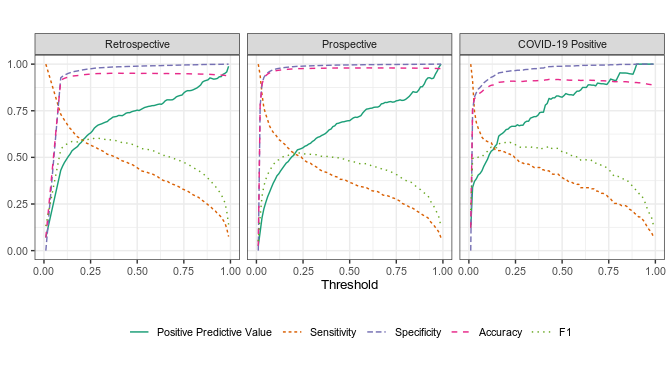

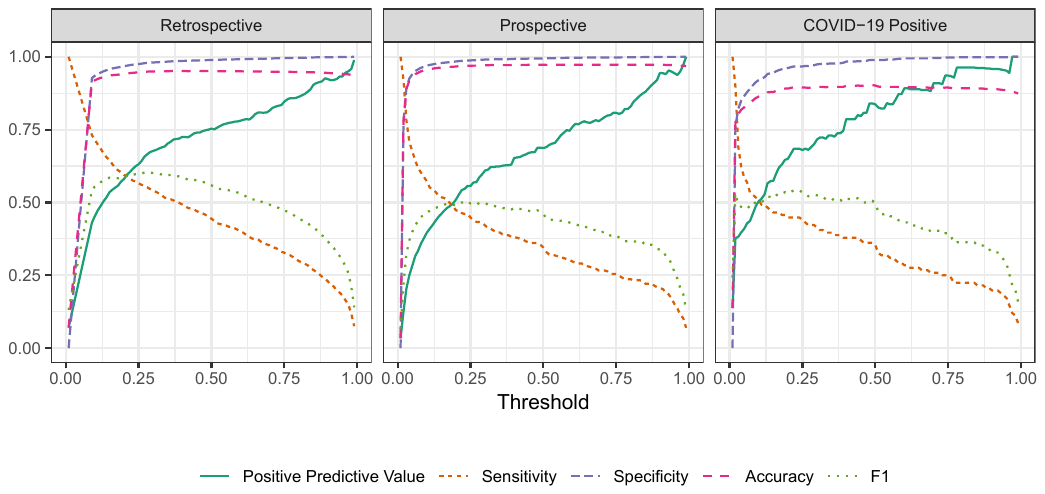

**eFigure 2: Confidence Intervals of Point-Wise Predicted Mortality in the Retrospective and Prospective Cohorts**

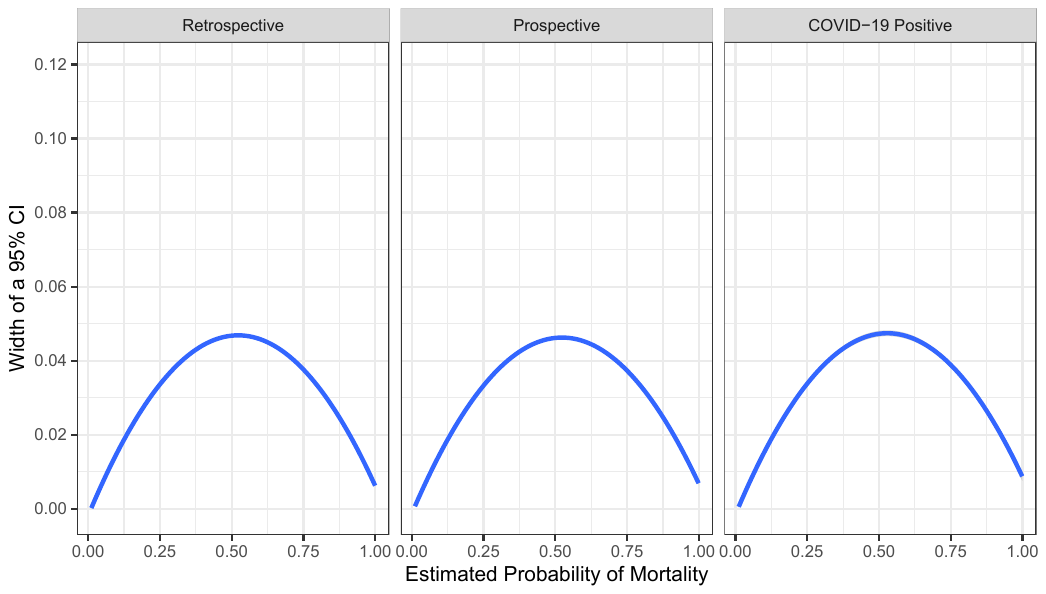

**eFigure 3: Average Predicted Mortality Stratified by Actual Mortality over the course of the Hospitalization in the Retrospective and Prospective Cohorts**

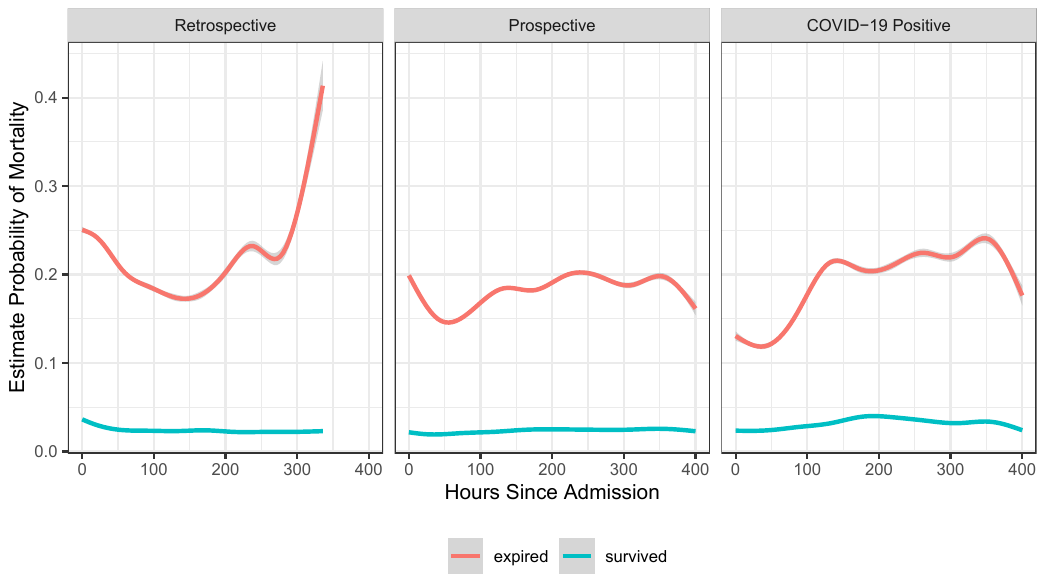
